# Rapid measurement and machine learning classification of color vision deficiency

**DOI:** 10.1101/2023.06.14.23291402

**Authors:** Jingyi He, Peter J. Bex, Jan Skerswetat

## Abstract

Color vision deficiencies (CVDs) indicate potential genetic variations and can be important biomarkers of acquired impairment in many neuro-ophthalmic diseases. However, CVDs are typically measured with insensitive or inefficient tools that are designed to classify dichromacy subtypes rather than track changes in sensitivity. We introduce FInD (Foraging Interactive D-prime), a novel computer-based, generalizable, rapid, self-administered vision assessment tool and applied it to color vision testing. This signal detection theory-based adaptive paradigm computes test stimulus intensity from d-prime analysis. Stimuli were chromatic gaussian blobs in dynamic luminance noise, and participants clicked on cells that contain chromatic blobs (detection) or blob pairs of differing colors (discrimination). Sensitivity and repeatability of FInD Color tasks were compared against HRR, FM100 hue tests in 19 color-normal and 18 color-atypical, age-matched observers. Rayleigh color match was completed as well. Detection and Discrimination thresholds were higher for atypical observers than for typical observers, with selective threshold elevations corresponding to unique CVD types. Classifications of CVD type and severity via unsupervised machine learning confirmed functional subtypes. FInD tasks reliably detect CVD and may serve as valuable tools in basic and clinical color vision science.

## Introduction

Conventional phenotypical categories for inherited color vision deficiency (CVD) are anomalous trichromats (AT), dichromats, and monochromacy, with mild to strong color vision defects, respectively. They can be further referred to as protan, deutan, or tritan types, with L-, M-, and S-cone relevant deficiencies, respectively(1). Identification and diagnosis of CVD are critical in many respects. In clinical applications, abnormal color vision may reveal genetic dyschromatopsia or early signs of neural pathway or systemic diseases(2). However, widely employed color vision tests are not ideal for detecting and monitoring acquired CVD progression or remediation, owing to their insensitivity, long testing time, difficulty of administration, and interpretation of their results.

Color matches conducted with an anomaloscope have been considered a gold standard for precise CVD diagnosis. An anomaloscope enables red-green Rayleigh matching and blue-green Moreland matching tests, in which the observer adjusts the mixed light side of the bipartite field to match the light in the reference side(3). However, anomaloscopes are expensive and a full examination of the color matches requires extensive instruction, expert administration, and exhaustive testing time. In addition, matching ranges of an extreme anomalous trichromat can be indistinguishable from those of dichromats. Pseudoisochromatic plates are capable of rapid screening and classification of CVD. Ishihara plates contain pseudoisochromatic numbers and curved lines, as well as vanishing plates with stimuli seen by only CVD patients. Hardy-Rand-Rittler (HRR) plates(4) exceed Ishihara plates, in that they contain universal color symbols instead of Arabic numbers, have a three-step severity scale for each CVD subtype, use a two-step psychometric protocol (i.e., what symbol do you see in which quadrant) to reduce probability of guessing, and are able to classify tritan(5). However, printed pseudoisochromatic plates have a number of shortcomings. They require a trained clinician to administer the test, and are generally insensitive to detect subtle changes of color detection due to the absence of a severity scale (Ishihara), a coarse severity scale (HRR), and the lack of cone-isolated colors. The Farnsworth-Munsell 100 hue test (FM100) provides a relatively complete color discrimination measurement for Protan, Deutan, and Tritan, by asking participants to arrange 85 caps according to color, but the task is extremely time-consuming, as is the analysis of FM100 error scores. These issues are addressed by D-15, an abridged version of FM100, however, interpretation of the error scores of both tests can be vague. CVD patients can also rely on luminance cues to pass the arrangement task or improve test scores(6).

Attempts to improve performance of traditional color vision tests have been made with computer-based tests. The Cambridge Color Test (CCT) adopted the pseudoisochromatic pattern from the above-described printed tests with a colored Landolt C orientation-identification stimulus embedded in static luminance noise, and utilizes forced-choice adaptive procedures(7). The Color Assessment and Diagnosis (CAD) test employs a moving chromatic square superimposed onto dynamic luminance noise(8). Resulting thresholds of both tests in vector length units can be used to classify the type and severity of CVD. In clinical contexts, testing duration is crucial. The CCT Trivector test shortened the testing procedure by assessing only the three confusion lines. Faster and simpler tablet-based tests which measure detectability along the three confusion lines were developed for better accessibility and for young children(9). The trade-off between testing speed and information collected has been the main cause of the disadvantages in these color vision tests.

In this article, we introduce and validate a generalizable, rapid, and self-administered computer-based procedure named FInD (Foraging Interactive D-prime), and adapt it to assess color detectability and discriminability. FInD uses an adaptive algorithm that measures visual performance thresholds for a wide range of visual functions, e.g. contrast sensitivity, motion- and form-coherence(10, 11). The algorithm selects stimulus strength derived from *d’*, a signal-to-noise ratio metric referring to the detectability of the stimulus, and calculates a new *d’* based on previous responses.

FInD Color Detection measures detection thresholds to L, M and S cone-isolating stimuli, reflecting the performance of individual cone types, while FInD Color Discrimination measures hue discrimination thresholds around 6 directions on a HSV (Hue, Saturation, Value) or equiluminant color plane. The two color spaces are used for different purposes – while equiluminant colors satisfy accurate measurement of color discrimination, HSV colors do not require careful calibrations so can be readily applied. The detection and discrimination tasks together interrogate an observer’s color vision system: the detection task classifies photoreceptor-level color sensitivity, and the discrimination task further quantifies the resolution of color perception. The computer-based Rabin Cone Contrast Test (RCCT) also adopted cone-isolating colors to screen CVD(12), in which color letters were presented without any background noise. FInD Color tasks instead use Gaussian blobs, therefore remove high frequency chromatic signals and avoid optotype familiarity. Dynamic luminance contrast noise is also added to mask potential luminance artifacts. Our first aim was to validate FInD Color detection and discrimination in terms of thresholds, testing duration and reliability in both color-normal (CN) and CVD groups. The second aim was to determine CVD type and severity using unsupervised machine learning (UML) classification with FInD Color detection and discrimination thresholds. The classification of CVD subtypes is difficult because genetic testing is expensive and laborious, and results of current behavioral tests for thresholds vary due to different stimulus types and tasks. Personalized threshold results generated with FInD enable the deployment of UML approaches to determine groups based purely on the behavioral performance of detectability and discriminability. The third aim was to compare FInD results against that of conventional clinical tools— HRR and FM100.

## Methods

The experiment protocol was approved by Northeastern University Institutional Review Board and followed the principles in the Declaration of Helsinki.

### Participants

19 participants (mean ± SD age: 26.2 ± 10.0; age range: 18-50; 9 females) with self-reported normal color vision and 18 (mean ± SD age: 23.1 ± 7.8; age range: 18-54; 1 female) with self-reported inherited color vision deficiency were recruited after providing informed consent and completing a demography and ocular history questionnaire. All participants had normal (20/20 or better) or corrected-to-normal visual acuity and had no history of eye diseases except for one participant with strabismus and two participants with amblyopia, among whom one amblyopia participant also had defective color vision. Two participants were diagnosed with attention deficit disorder and depression, respectively, and both received medical treatments.

### Apparatus

Experimental procedures were programmed by Psychtoolbox(13) in MATLAB (MathWorks, Natick, MA), and presented on a 32” 4K LG display with a resolution of 3840×2160. The display was gamma-corrected with SpyderX elite colorimeter (Datacolor, Lawrenceville, NJ) and the spectra and luminance were measured with a Photo Research PR-650 spectroradiometer (Photo Research, Chatsworth, CA). Luminance of the mid-grey background was 90.3 cd/m^2^. Participants viewed the screen binocularly at a distance of 111 cm, subtending a visual angle of 35 deg × 20 deg, with the head position stabilized in a chin rest. Standard illumination applied for HRR and FM100 administration was provided by a Sol•Source daylight lamp (117V, 50/60 Hz) manufactured by GretagMacbeth. The time was recorded using a standard mobile phone timer application or by the computer for FInD tests.

### Tasks and Stimuli

Four methods were compared in this study: Hardy-Rand-Rittler (HRR) Pseudoisochromatic Plates (4th Edition), Farnsworth-Munsell 100 hue test (FM100), Rayleigh color match, and FInD Color Detection and Discrimination tasks.

### Hardy-Rand-Rittler (HRR) Pseudoisochromatic Plates

HRR was conducted by the experimenter flipping the plates and participants reporting the shape and location of test color symbols under controlled lighting.

### Farnsworth-Munsell 100 hue test (FM100)

Participants were asked to arrange the 85 caps according to reference colors in each testing case. The four cases were completed in random order.

### Rayleigh Color Match

Rayleigh color matching data were collected on 14/19 CN and 6/18 CVD with an Oculus HMC Anomaloscope (Oculus, Germany). Participants were asked to complete 8 measurements (4 with each eye) and an additional matching range identification trial.

### FInD Color detection

Stimuli utilized in FInD Color Detection task were cone-isolated Gaussian blobs (σ=1°, support diameter = 4°) (Figure 4a left). L-, M-, and S-cone isolating directions in RGB unit were calculated by integrating Stockman-Sharpe cone fundamentals(14, 15) and the measured display spectra, then weighted by cone contrasts (detailed computation steps can be found in He, Taveras-Cruz (16) Appendix; see Supplementary materials for exact values used). Stimulus contrast was adaptively controlled by the FInD algorithm.

### FInD Color discrimination

FInD Color Discrimination task measures discriminability of a pair of two small Gaussian blobs (σ=0.6°, support diameter=3.6°) with different colors (Figure 4a right). Each stimulus contains a pair of colors that were selected from the HSV color space (Figure 4b). Six hues (H = red, yellow, green, cyan, blue, magenta), 3 primary and 3 confusion axes, in the half-brightness (V=0.5) HSV plane were tested separately with two saturation levels (S = 100% or 50%). For each selected hue axis, two stimulus colors were selected at the same angular distance away from this hue axis in opposite directions. The angular distance between test colors in HSV space was adaptively controlled by the FInD algorithm.

### Procedures

Each participant completed the four tasks once (test session) or twice (test-retest sessions) in random order after optometric screening.

The FInD adaptive algorithm measures *d’* to efficiently estimate thresholds. In short, *d’* is a measure of detectability or discriminability in signal detection theory. As in Figure 4c, *d’* is the distance between the noise and signal distribution means, and the location of the criterion (λ) directly affects the proportion of responses corresponding to “hit”, “miss”, “false alarm”, and “correct rejection”. The initial stimulus contrast (detection) or angle distance (discrimination) was determined by pre-defined approximate estimates of slope and threshold to derive the test range to span difficult (*d’*=0.1) to easy (*d’*=4.5) stimuli. The independent variable of the stimuli for the first chart were sampled in log steps across this test range. Subsequent charts use the combined responses across all previous charts to re-estimate *d’* (Figure 4e).

Stimuli were presented in charts (Figure 4a) containing 4×4 cells (6°×6°) at the center of the screen with each cell containing one stimulus at pre-selected contrasts (detection) or hue difference (discrimination) embedded in 8 Hz dynamic luminance noise, with check size 10 arcmin and ±20% luminance contrast. A high stimulus-intensity example and instructions were provided at the upper left corner of the screen to help participants identify the targets to be foraged. Participants were informed that target stimuli were present in some but not all cells, and the number of targets varied from chart to chart. They were instructed to click on any cells that contained “faint versions” of the stimulus (detection) or on cells that contained blob pairs that have different colors (discrimination). The chart stayed on the screen until the observer completed the current chart by clicking on the exemplar. *d’* was then calculated based on hit/ miss/ correct rejection/ false alarm classifications as a function of stimulus contrast (detection) or hue difference (discrimination). The estimate of *d’* from all previous responses was used to generate the stimulus range (contrast or hue difference) in the next chart. A series of three charts for each stimulus were presented in interleaved order (Figure 4d).

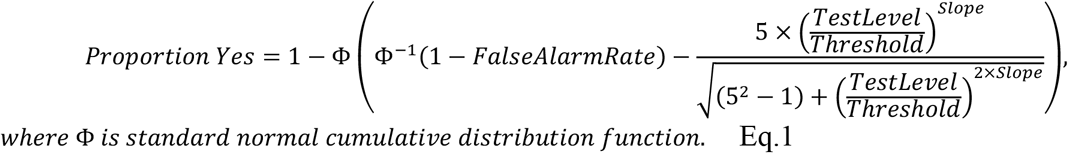

## Results

Data analyses were performed in MATLAB (MathWorks, Natick, MA). Descriptive statistics of FM100 and FInD tasks for the CN group are summarized in Table S1. The analyses below are based only on the test session data except for the repeatability analysis.

### HRR

All CN observers passed the first ten diagnostic plates of HRR. The time recorded for 12 CNs to pass was 37 seconds on average. All CVDs self-reported to have color vision deficits failed HRR and were classified as protan or deutan with mild, medium, or strong defect (Table 1), and time spent for 13 CVDs was 2’53” on average to complete all 24 plates of HRR. One CVD observer with mild red-green deficiency achieved equal protan and deutan scores thus could not be classified.

**Table 1.**
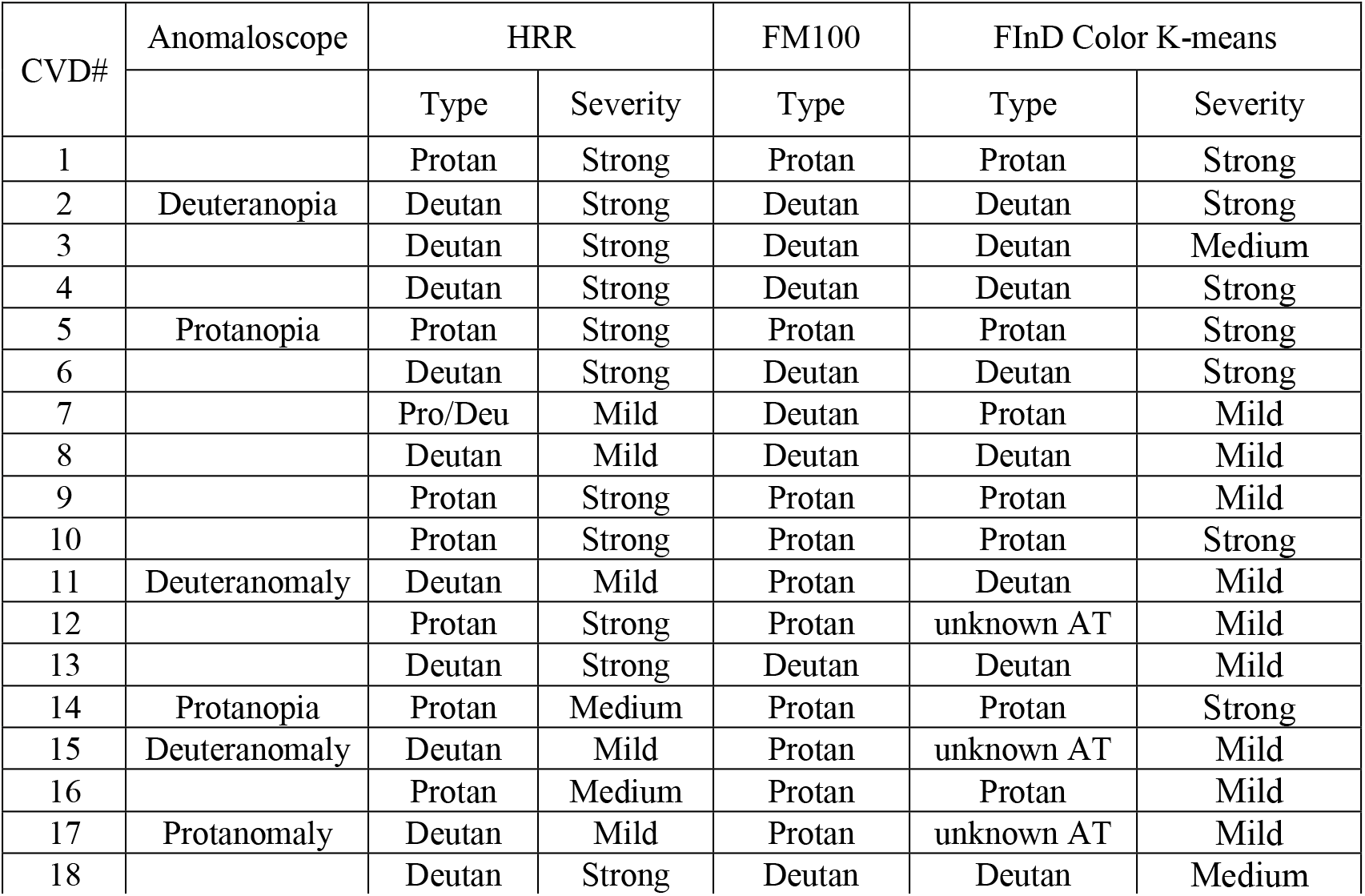
Identification of CVD type and severity for 18 CVD observers for the three methods.

### FM100

Total error scores (TES) and right-half mid-point (MP) were reported for FM100. TES is calculated as the sum of the error scores for each color with 2 subtracted from each error score. For CNs, TES is expected to range from 0 to 100 where superior, average, and low discrimination abilities are indicated by error scores ranging from 0∼16, 20∼100, and more than 100. In our sample 42%, 53%, and 5% of the CN observers respectively fell in the three categories (Table 1). Mean and standard deviation of the TES of the CN group are reported in Table S1, and average error score pattern is shown in the left panel of Figure 1 for the CN group. Note the radial axis range, where the center of the error score pattern is 2, indicating perfect responses, with the largest error score scale being 3.5. Our TESs for CNs (2.42±0.24) are comparable to those reported in Knoblauch, Saunders (17). Error score pattern of one CVD participant (CVD#5) is shown in the right panel (note radial axis range is 2-16). Patterns for each CVD observer are provided in Figure S1. TES of 11% CVD observers exceeded the error score range of our CN group while 89% CVDs have TES larger than 100. The right-half MP contains the median of error scores of cap sequence 43 to 84, and was used to identify type of defect. The classification criteria are indicated in the right panel of Figure 1 according to the FM100 manual: protans, deutans, and tritans have right-half mid-points in ranges 62∼70, 56∼61, or 46∼52, respectively. TES and MP scores of the CVD group are reported in Table S2. Average time taken for testing participants were 13’32” for 18 CNs and 13’23” for 17 CVDs. As for plotting the pattern and calculating TES and MP manually, 3’17” for CNs and 14’48” for CVDs were taken on average. Computer-based data analysis algorithms can largely improve efficiency of FM100 test by reducing the data processing time.

**Figure 1.**
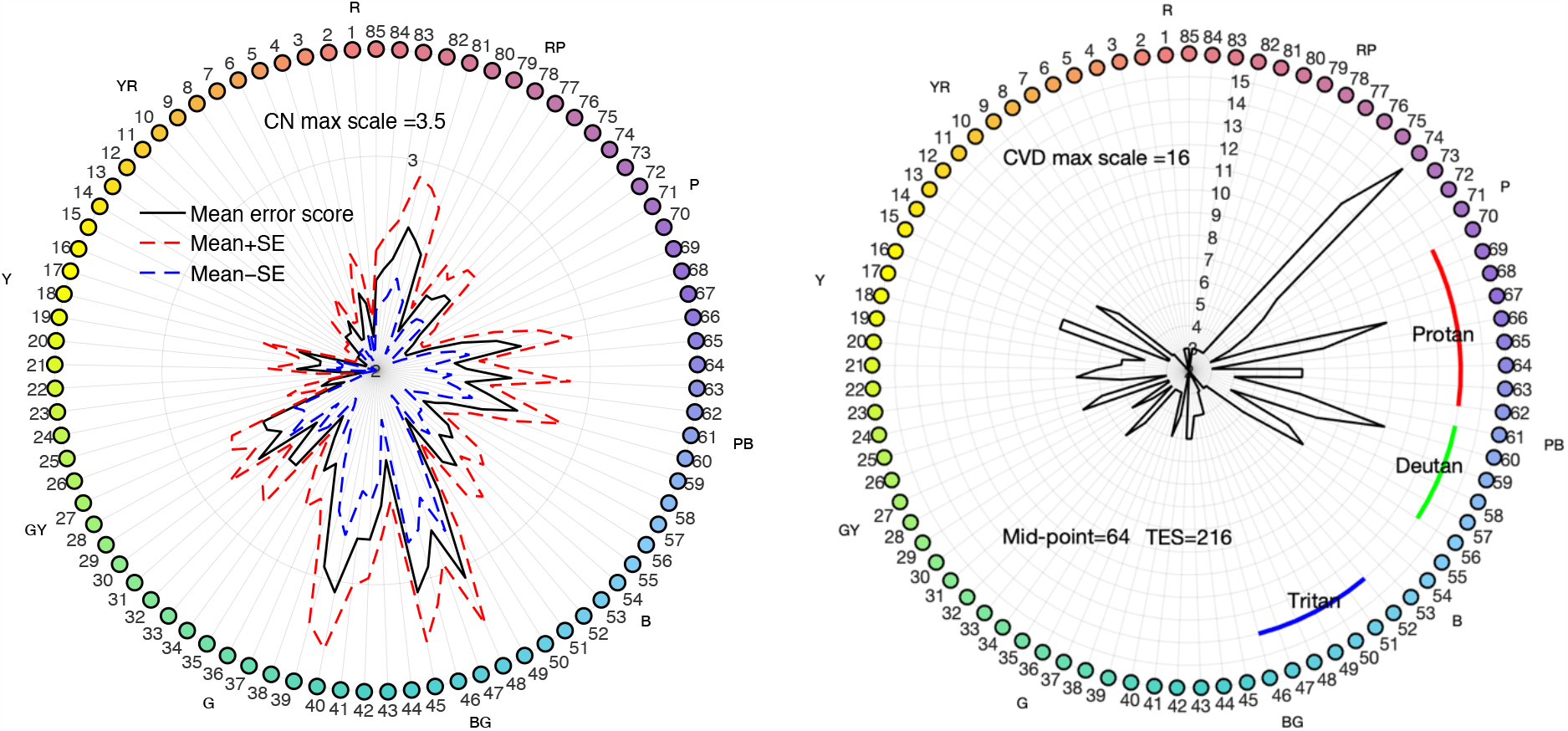
FM100 hue test results. Left: average error score pattern of 19 CNs. Hues of colored caps are numbered from 1 to 85 with the corresponding mean error score (black line) and standard error range indicated along radial coordinates for each hue. Upper and lower standard error ranges are depicted by the red and blue dashed lines, respectively. The outermost circle where the color dots reside represents an error score of 3.5, and the center error score is 2, indicating the lowest possible error score. Right: error score pattern (black line) of an example CVD observer (CVD#5). Note that the largest radial scale is 16. Mid-point and TES of this observer as well as diagnostic curves (color arcs) are also shown. Mid-point of this observer falls in the protan range.

### Rayleigh Color Match

Testing time duration for Rayleigh match on an anomaloscope was 10.64 min on average for 14 CNs to complete 8 measurements (4 with each eye) and 15.07 min for 6 CVDs to complete all 8 measurements with an additional matching range identification trial. Detailed testing procedures and results are reported in the Supplementary material. All 14 CN participants produced normal anomalous quotients and all 8 CVDs produced atypical quotients. 6 CVD classifications agreed with HRR classification and 4 agreed with FM100 classifications (Table 1).

### FInD Color detection and discrimination thresholds and K-means clustering

FInD Color detection thresholds for each CVD observer are shown in the top panels in Figure 2, and low-saturation discrimination thresholds are shown in bottom panels (Figure 2b; high-saturation discrimination thresholds are shown in Figure S2). For both tasks, data of CNs tend to cluster at lower thresholds and have relatively small variance, whereas there are large individual differences among CVD observers with different patterns and degrees of selective threshold elevation likely corresponding to CVD types. Average (±standard deviation) time durations taken for CN and CVD participants to complete all trials in the detection task were 4’57” (±1’59”) and 5’08” (±1’45”), and were 19’57” (±5’33”) and 18’04” (±5’32”) to complete all trials in the discrimination task, respectively. Given that two saturation levels were investigated in the discrimination task, one test level takes only half of the time reported (less than 10 min).

**Figure 2.**
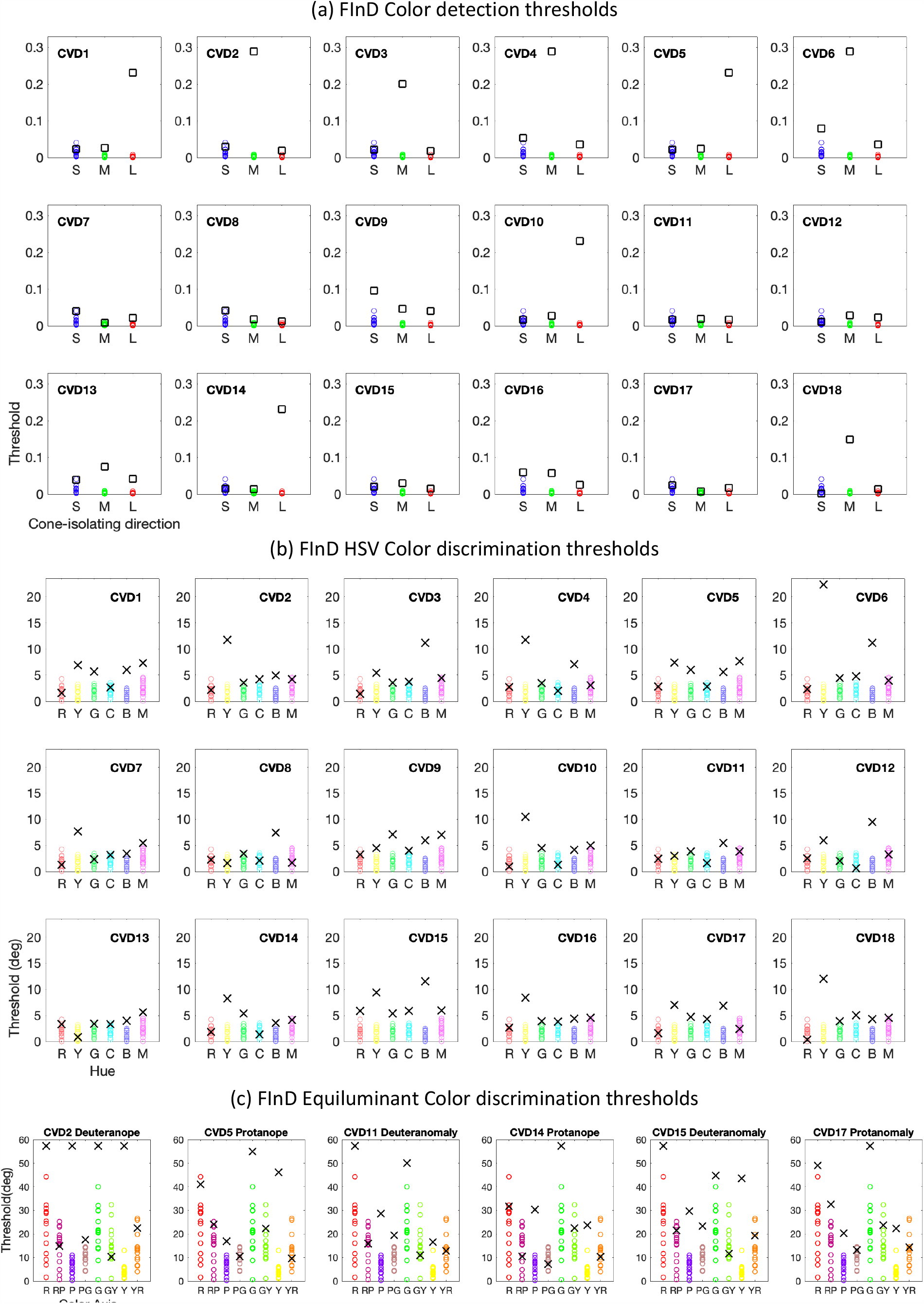
FInD Color detection and discrimination thresholds. Thresholds of all CN participants are plotted as colored circles in all panels as references, and thresholds of each CVD observer are denoted by black squares (detection) or crosses (discrimination) in separate panels. (a) FInD Color detection thresholds (upper panels) are plotted as cone contrast vector length and (b) low-saturation discrimination (lower panels) thresholds are plotted in degrees of HSV color space angle. (c) shows results of 6 CVD participants for the FInD Color discrimination task with equiluminant, colored stimuli.

To set up diagnostic criteria for FInD Color results, we sought to achieve an automated classification of CVD type and severity using unsupervised machine learning (UML). To that end, a K-means clustering algorithm was applied with inputs being subsets of the detection and discrimination threshold datasets. A two-step classification was performed (see details in Supplementary section: *K-means classification*). The first step took LM detection thresholds as inputs and was able to segregate a large group. Observers in this group include CN and potential anomalous trichromats (AT) with low LM thresholds (Figure 3a green circles). We refer to this group of participants who have defective color vision but low detection thresholds as ‘potential AT’ for simplicity in the following paragraphs. Three smaller clusters were also identified, one with high L and low M thresholds (CVD#1,5,14; likely protanopes; Figure 3a blue diamonds), one with high M and low L thresholds (CVD#2,4,6; likely deuteranopes; Figure 3a cyan squares), and one with intermediate M and low L thresholds (CVD#3,18; likely extreme deuteranomaly or deuteranopes; Figure 3a orange triangles). Observers who were classified in the three small groups received consistent classifications as from HRR, FM100, and anomaloscope data if available (Table 1). The second classification step took LMS detection thresholds and low-saturation yellow (Y), blue (B) and magenta (G) hue discrimination thresholds as inputs, and divided the large group from the first step to four smaller clusters: one CN group (Figure 3b-f: “1”s surrounded by green circles), one deutan group (likely deuteranomaly; Figure 3 b-f: “3”s surrounded by red squares; 2 received consistent HRR and FM100 classifications, and 1 received inconsistent HRR and FM100 classifications but was classified as deuteranomaly by the anomaloscope), one protan group (likely protanomaly; Figure 3 b-f: “4”s surrounded by red squares; 2 received consistent HRR and FM100 classifications, and 1 received inconsistent HRR and FM100 classifications) and one unknown AT group (Figure 3 b-f: “2”s surrounded by red squares; 2 received inconsistent HRR and FM100 classifications, and were classified as protanomaly and deuteranomaly, respectively, by the anomaloscope, and 1 received consistent HRR and FM100 classifications as protan). With the lack of genetic measurements available for the participants, however, we are not able to relate the subgroups to any specific genotype.

**Figure 3.**
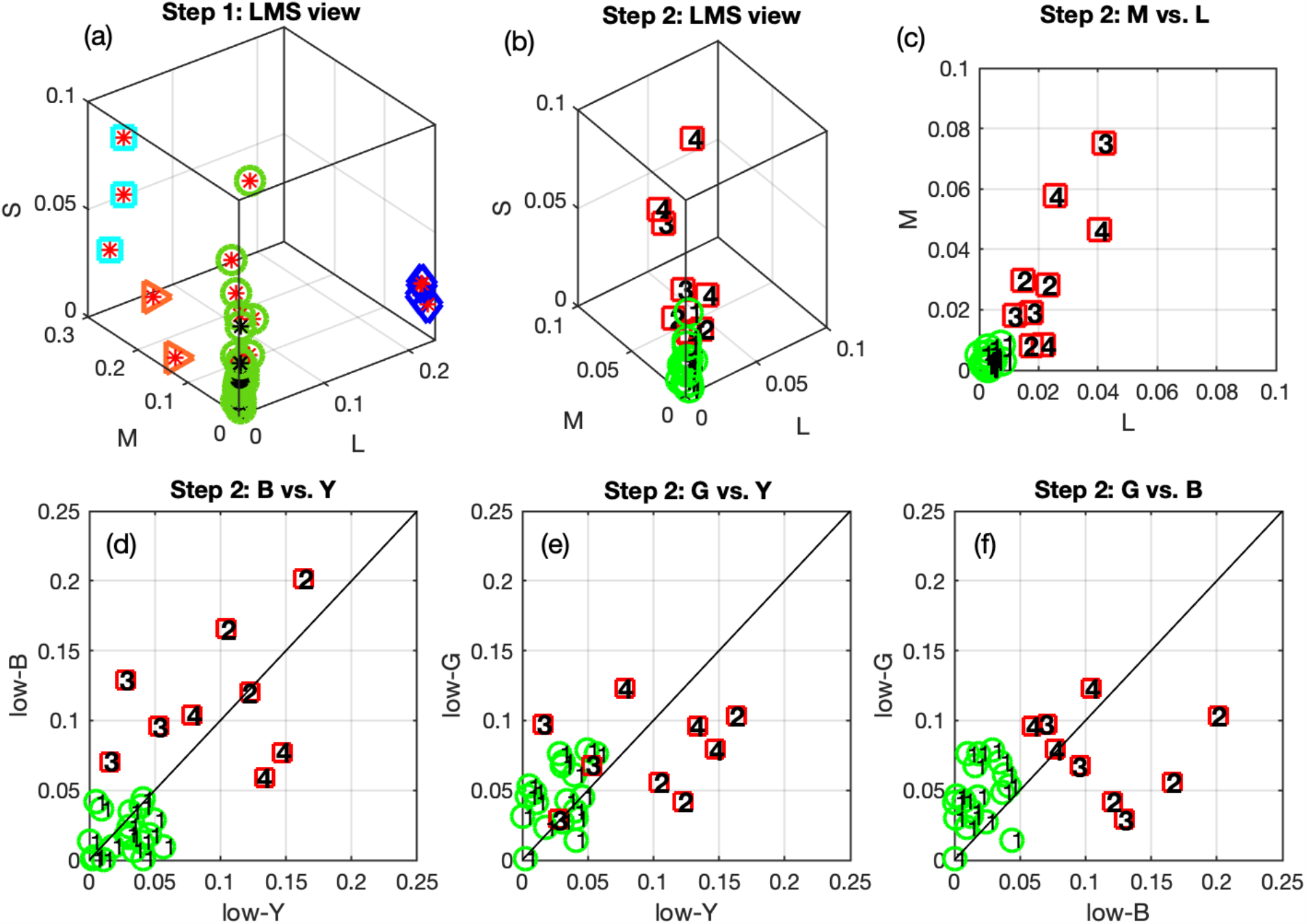
Classification of CN and CVD results. (a) Step one clustering results illustrated in LMS detection threshold space. All 37 individual data points are shown. CVD and CN individuals are represented by red and black asterisks, respectively. Four clusters, denoted in differently shaped and colored symbols (green circles, blue diamonds, cyan squares, and orange triangles) around the asterisks, are identified. (b)-(f) show step two clustering results. Only individual points surrounded by green circles (n=28) in (a) are taken and plotted. These thresholds were clustered as four groups denoted by numbers. Red squares and green circles represent CVD and CN, respectively.

**Figures 4.**
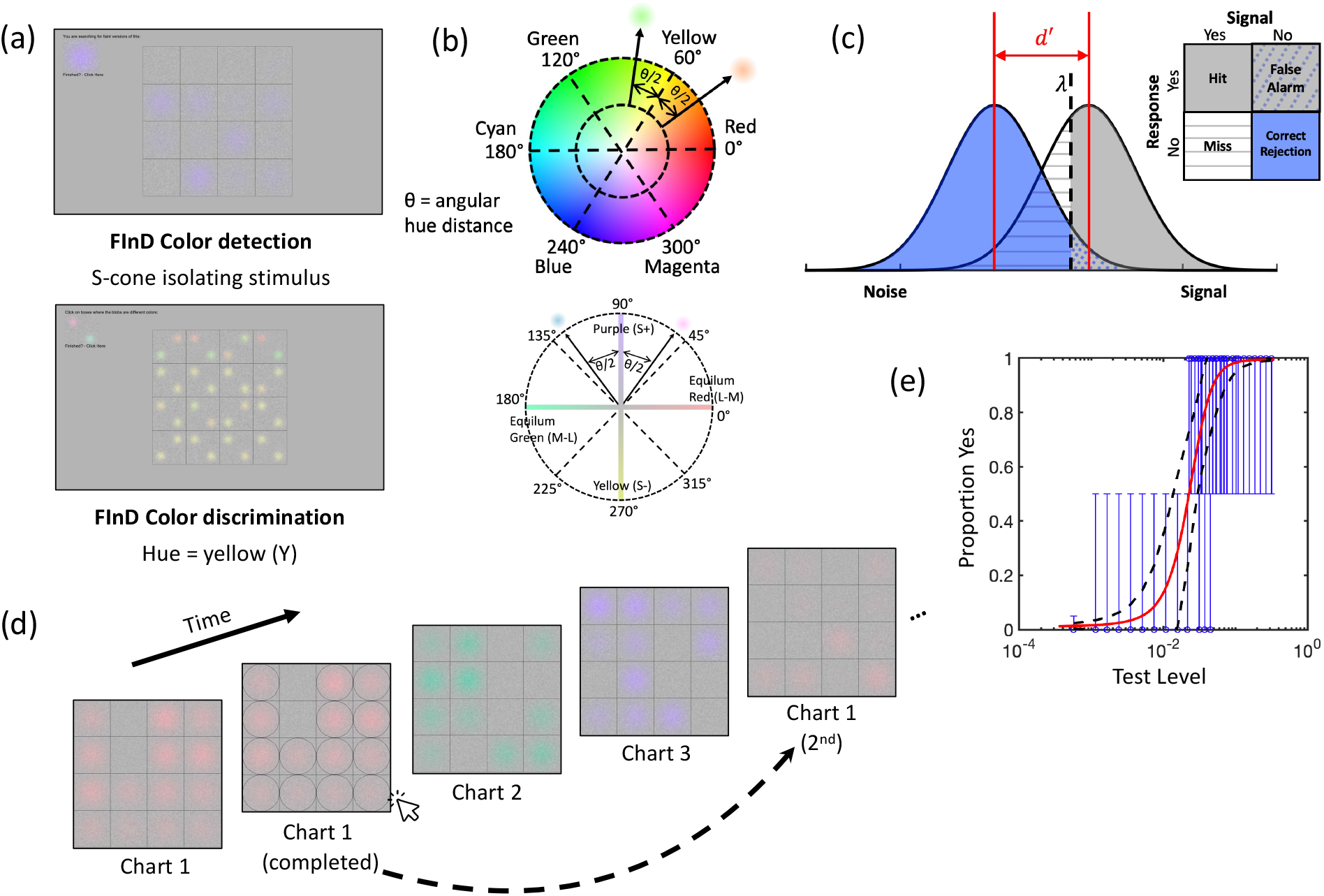
Illustration of FInD stimuli and experimental procedures. (a) FInD detection (top) and discrimination (bottom) task interfaces. (b) The top color wheel shows a cross-section of the HSV space for V=1, from which six hues (0° to 300° in 60° steps) and two saturation levels (0.5 and 1) were chosen and used in the discrimination task. If, for instance, yellow (60°) discrimination were tested, two colors are symmetrically selected the same angular hue distance (θ/2) away from yellow with a fixed saturation level. The bottom color wheel shows the equiluminance plane with four primary axes representing the red-green and blue-yellow postreceptoral mechanisms (c) Illustration of signal detection theory. The noise distribution (blue) and signal distribution (grey) bell curves lie on the normalized Z-score abscissa. Detectability or discriminability (*d’*) and criterion (λ) are depicted. The areas under the curves correspond to “hit”, “miss”, “false alarm”, and “correct rejection”, respectively, according to stimulus presentation and response. (d) FInD experimental procedures with cone isolating direction detection stimuli as an example. The dashed arrow represents the adaptive procedure that selects a range of stimulus intensities on the 2^nd^ chart based on analysis of the responses to stimuli on the 1^st^ chart. (e) An example of a typical psychometric function: blue data show the probability that the observer reported the presence of a stimulus as a function of intensity, vertical lines show binomial standard deviation, red curve shows the best fitting function for Eq.1, and black dashed lines represent upper and lower 95% confidence intervals. The separate data point on the left of other data points indicates the false alarm rate.

### Test-retest reliability

Thirteen CN and eight CVD participants completed the retest session. All CNs passed HRR on both tests. HRR categorization of two CVD observers were changed: one observer (CVD#14) stayed the same type (protan) but with a different severity (medium to strong), and the other observer (CVD#12) changed in both type and severity (strong protan to medium deutan). Bland-Altman analyses(18) show no significant learning effect and bias (see details in the Supplementary material).

### Comparison of the methods

Classification results with three methods for the 18 CVD observers are compared in Table 1. As we used consistent Rayleigh match, HRR and FM100 CVD type classification as references in the two-step unsupervised classification, agreement between FInD Color and the other tasks should be expected. For the observers classified as strong or medium protan or deutan (likely protanope or deutanope) using FInD Color LM detection thresholds, CVD type classification of the three methods agree perfectly (CVD#1,2,3,4,5,6,14,18). However, LM detection thresholds alone were not able to distinguish between CN and AT as they both have relatively low cone specific detection thresholds. With the addition of the hue discrimination thresholds, the algorithm successfully distinguished CN observers from anomalous trichromats (AT), and further assign potential AT to CVD subtypes. The second-step classification results agree for all 19 CN observers and 5 out of 9 AT observers. The remaining four AT observers (CVD#7,11,15,17) received inconsistent diagnoses from HRR and FM100, therefore for FInD classification, we assign them to the CVD categories in which the members in their clusters fall, which necessarily agree with one of their HRR or FM100 diagnoses, but not both. It’s worth noting that in one cluster (CVD#:12,15,17), although one of the participants (CVD#12) received consistent HRR and FM100 diagnoses in the test session, the retest HRR diagnosis changed type, so all three participants received confusing HRR and FM100 diagnosis when the retest session results are considered. This cluster is then referred to as the unknown AT group. As for testing durations, a FInD Color detection task with three testing directions and three trials per direction (CN: 4’57”, CVD: 5’08”) took longer than HRR (CN: 37”, CVD: 2’53”), but quick screening with only one trial took shorter (1’43”) for CVD. A single level FInD Color discrimination task (CN: 9’59”, CVD: 9’02”) was more rapid than FM100 (CN: 13’32”, CVD: 13’23”). FM100 testing can be further prolonged due to the additional time spent on data processing.

## Discussion

The current study introduced and measured performance of the newly designed FInD Color detection and discrimination tasks, and compared them against HRR, FM100 hue and Rayleigh match tests. FInD is rapid, self-administered, and easy to use, without strict operating regulations other than the normal usage of a computer, so can be readily grasped by patients or children(19).

The combination of color detection and color discrimination performance also provides richer information about CVD color perception, and enables classification of color vision subtypes using unsupervised machine learning (UML) classification. In comparison, HRR, although rapid and easy, provides limited information about atypical color perception, and is not able to classify CVD type for very mild cases (CVD#7). FM100 measures color discrimination but does not have strict and precise diagnostic metrics. Even in such a small sample, some CVD observers obtained lower error scores than observers with normal color vision but poor discrimination. Moreover, the actual error score pattern shows large degree of variation instead of a clearly classifiable pattern. FInD Color tasks afford a remedy for these shortcomings, which may be critical for detecting and tracking progression or remediation of acquired CVD in neuro-ophthalmic disease(2). Furthermore, FInD discrimination task significantly reduces testing and analysis duration compared to FM100, and FInD detection task, although taking similar amount of time compared to HRR for CVD testing, is more informative.

In the present implementation of FInD Color detection, we calibrated the monitor to generate and test detection thresholds for short-, medium- and long-wavelength sensitive cone-isolating stimuli, based on standard cone fundamentals (see Methods). For FInD Color discrimination, we employed HSV color space to generate stimulus pairs of differing colors. HSV is based on subjective estimates of distance in perceptual color space, which is convenient for measurement of color perception on standard computer displays. However, different color hues in HSV may not be equiluminant, and therefore participants could utilize luminance differences instead of color differences to select cells that contained blobs of differing color. We attempted to mask any such cues with 20% contrast dynamic luminance noise, and in the Supplementary materials we report the magnitude of luminance artifacts in our display. The results show that artifacts differ with test hue angle in a pattern that is inconsistent with differences in discrimination threshold and are less than 6 cd/m^2^ for all test axes at the highest discrimination difference for CN participants. A separate luminance-matched achromatic blob discrimination task was conducted to evaluate magnitude of luminance artifacts, and thresholds for luminance difference are reported in Table S7. Sensitivities of CN observers to low-saturation luminance differences never exceed the artifact generated at a threshold level of 5° hue angle. However, Sensitivities to high-saturation luminance differences along three hue axes (R, C, G) exceed the threshold at 5° hue angle. In this case, the results of the FInD Color discrimination test are conservative, and we would expect color discrimination thresholds to be even higher for CVD participants. To assess performance of the discrimination task in a color space free of luminance artefacts, a subset of participants (6 CVDs and 14 CNs) were available to complete the discrimination task but with colors chosen from an equiluminant plane (See Figure 2(c), and details in Supplementary, *FInD Color discrimination task with an equiluminant color plane*). Eight color axes were measured. Results show that response patterns using the equiluminant plane are similar to that of the HSV space: both protans and deutans have significantly elevated thresholds at yellow, green, blue/purple axes, with thresholds for red and other intermediate axes being more variable (see p-value tables in Supplementary).

Widely used CVD categories adopted by many color vision screening tests were predicated on color vision phenotype which allows rapid detection of abnormal performance. Albeit simple, the conventional categories disguise the variable nature of CVD. The well-known large individual variations in CVD make the diagnostic groups less distinguishable, and their relationship to genotypes remains unclear(20, 21). For instance, in addition to the protanomaly category, one CVD type (Pseudo-protanomaly), while having LM photopigment peak similar to protanopes, is able to produce trichromacy with differed optical density(22). The story can become more complicated when interactions between postreceptoral signals are considered. Brain plasticity seems to elicit compensatory adjustments so that the actual perceptual color ability loss for CVD can be reduced, further blurring boundaries between discrete phenotypes(23). UML has the ability to group individuals with similar features to the same category, and has previously been used to successfully identify subtypes of other diseases(24). Finding categories for groups is a non-trivial challenge for the following reasons: using prior information such as self-report of CVD or clinical results to classify data is dependent on the methods used in the clinical tests; Furthermore, treating CN and CVDs as one distribution and using multiple standard deviations away from the mean of CNs does not consider the known CVD subtypes that belong in separate distributions. Hence, we decided to use UML technique — k-means to address these problems to classify groups without assigning a group *a priori*. The novel application of using k-means clustering algorithm to establish CVD classification criteria in the present work, sheds light on the competence of machine learning techniques in capturing obscure response patterns in psychophysical measurements of typical and atypical color vision, thus possessing the potential to reveal the continuous and variable nature of CVD. As a result, the CVD group can be better categorized into subtypes, and inconclusive CVD cases that received confusing diagnoses from traditional tests can be better understood and treated.

The addition of hue discrimination thresholds in the second-step classification was essential to discern potential ATs, as detection thresholds alone did not effectively differentiate potential anomalous trichromats from normal trichromats (Note that the LMS cone isolating directions used in the detection task are computed based on a standard observer with normal color vision, therefore certain degree of individual differences should always be considered). This finding suggests complexity in AT phenotypes, echoing the previous evidence(20). We caution that inferences about AT features made based upon current results should remain provisional until larger datasets are acquired, which would also benefit the comprehensive evaluation of the FInD method. As discussed above, even though the HSV plane in the discrimination task is not equiluminant, our classification results demonstrate its ability to assay response pattern differences between CN and CVD, with significantly different discrimination patterns observed and the pattern of findings was the same for an equiluminant color space. Apart from HSV, the FInD tasks implemented in this study required careful display calibration to generate cone-isolating directions (for an ideal observer model) and equiluminant color planes. The same concerns prevail for the anomaloscope and for the light source and pigment decay of HRR and FM100 tests. Therefore, deployment of these approaches will depend on calibration of the use of specific model specifications. The ability of FInD Color tasks to detect and classify tritan deficiencies, inherited or acquired, was not tested in the current study. Future efforts are needed to verify whether the unused features, S detection and the rest of the hue discrimination thresholds, are critical for the categorization of tritans.

In summary, this proof-of-concept study has shown that FInD Color tasks can provide continuous color detection and discrimination threshold estimates that may track change in CVD and could therefore serve as a rapid and easy-to-use tool for clinical monitoring and diagnosis. The HSV colors are equally diagnostic compared to the equiluminant colors. FInD also has potential in basic color vision research, in that the combination of FInD Color tasks with UML technique might provide insights about hidden structures in the data and further assist the understanding of defective color mechanisms.

## Data Availability

Data produced in the present study are available upon reasonable request to the authors.

## Acknowledgements

The work is supported by NIH Grant R01 EY029713.

