## Supplementary for "Rapid measurement and machine learning classification of color vision deficiency"

### Supplementary Materials

#### *Tables and figures*

| CN group | | FM100 TES | FInD Detection | | | FInD Discrimination (deg) | | | | | |
|---|---|---|---|---|---|---|---|---|---|---|---|
| | | | S | M | L | R | Y | G | C | B | M |
| Test (n=19) | | | | | | | | | | | |
| Mean (STD) | Low saturation | 35.79 (31.73) | 0.0124 (0.0093) | 0.0031 (0.0021) | 0.0031 (0.0018) | 0.0307 (0.0186) | 0.0273 (0.0176) | 0.0326 (0.0152) | 0.0323 (0.0168) | 0.0189 (0.0146) | 0.0458 (0.0220) |
| | High saturation | | | | | 0.0100 (0.0064) | 0.0122 (0.0085) | 0.0210 (0.0129) | 0.0224 (0.0145) | 0.0123 (0.0089) | 0.0240 (0.0190) |
| Retest (n=13) | | | | | | | | | | | |
| Mean (STD) | Low saturation | 18.46 (13.12) | 0.0105 (0.0056) | 0.0015 (0.0008) | 0.0028 (0.0013) | 0.0295 (0.0137) | 0.0155 (0.0140) | 0.0317 (0.0145) | 0.0346 (0.0197) | 0.0164 (0.0163) | 0.0453 (0.0265) |
| | High saturation | | | | | 0.0090 (0.0078) | 0.0133 (0.0085) | 0.0253 (0.0172) | 0.0205 (0.0126) | 0.0116 (0.0075) | 0.0349 (0.0198) |

Table S1. Descriptive statistics of test and retest sessions for the color normal (CN) group.

| CVD# | Total error score (TES) | | Mid-point (MP) | |
|---|---|---|---|---|
| | Test | Retest | Test | Retest |
| 1 | 348 | - | 62 | - |
| 2 | 244 | - | 60 | - |
| 3 | 184 | - | 61 | - |
| 4 | 184 | - | 60 | - |
| 5 | 216 | - | 64 | - |
| 6 | 172 | 132 | 59 | 64 |
| 7 | 204 | 140 | 60 | 66 |
| 8 | 192 | - | 60 | - |
| 9 | 128 | 148 | 65 | 64 |
| 10 | 180 | - | 67 | - |
| 11 | 56 | 72 | 71 | 71 |
| 12 | 128 | 136 | 67 | 63 |
| 13 | 180 | - | 60 | - |
| 14 | 180 | 152 | 67 | 64 |
| 15 | 168 | 112 | 64 | 66 |

| | | | | |
|---|---|---|---|---|
| 16 | 112 | - | 68 | - |
| 17 | 100 | 92 | 71 | 69 |
| 18 | 144 | - | 60 | - |

Table S2. Farnsworth-Munsell 100 hue test (FM100) total error score (TES) and right-side mid-point (MP) of test (n=18) and retest (n=8) sessions for color vision deficiency (CVD) observers.

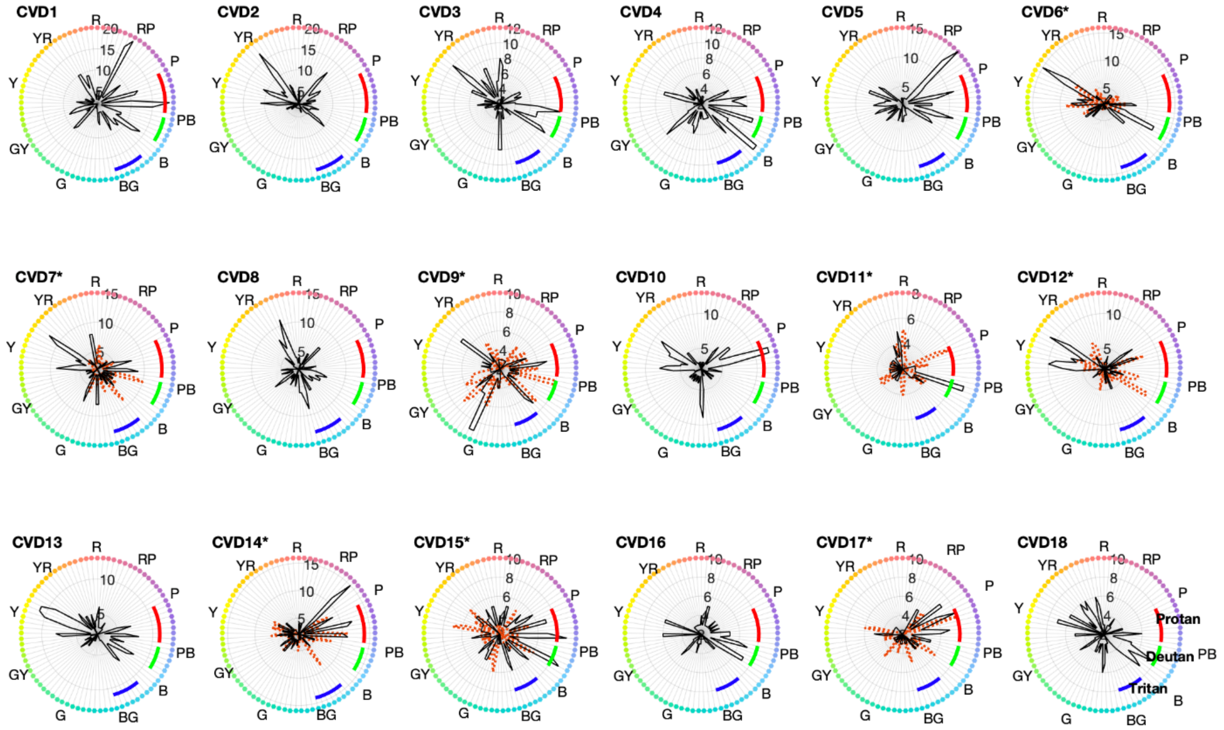


Figure S1. FM100 error score pattern for 18 CVD observers. Black solid lines and orange dotted lines represent test and retest session scores, respectively, for each color cap, with the corresponding hues denoted as color dots at the outermost circle and labeled as: R (red), YR (yellow-red), Y (yellow), GY (green-yellow), G (green), BG (blue-green), B (blue), PB (purple-blue), P (purple), and RP (red-purple). Radial scales which represent error scores are not consistent across participants for better visualization of the pattern. Diagnostic criteria for protan, deutan, and tritan are denoted as red, green, and blue curve segments, respectively. Participants who completed the retest session are indicated by * after the numbering. Error score pattern of CVD5 is also shown in Figure 1 right panel.

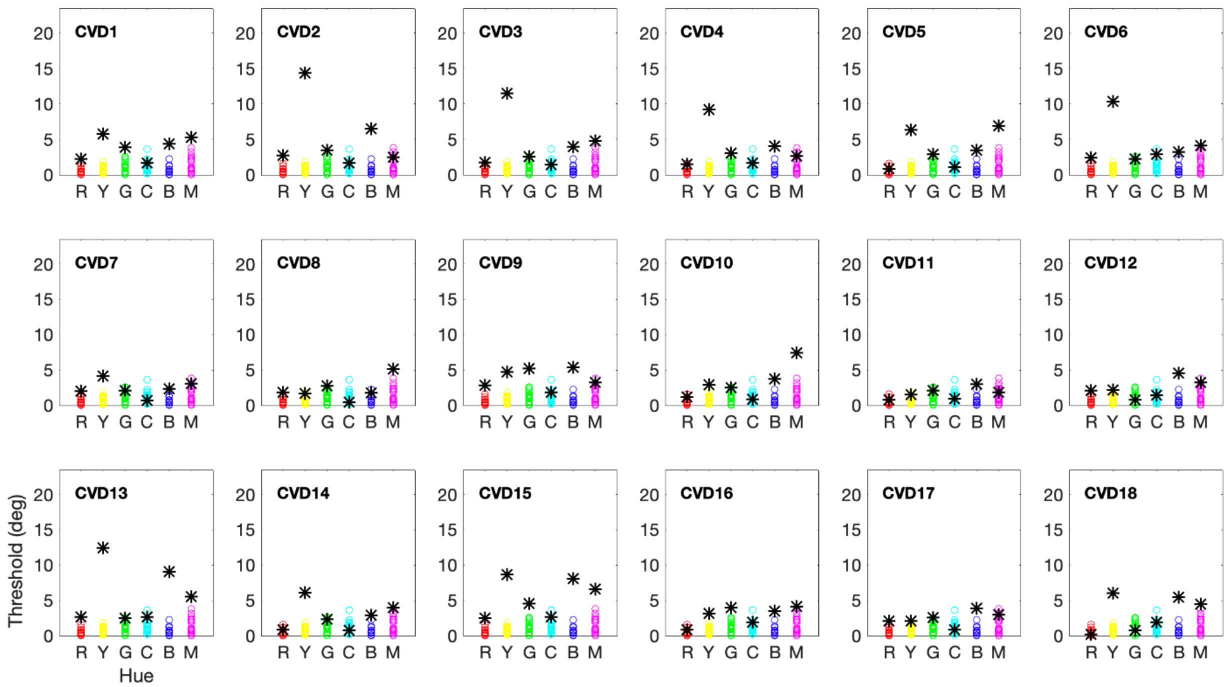


Figure S2. FInD Color high-saturation discrimination thresholds in degree. Thresholds of CN observers are represented in colored circles in every panel as references. Thresholds of the 18 CVD observers are represented as black asterisks.

#### ***K-means classification***

The K-means clustering algorithm used for determining FInD Color criteria was the MATLAB function *kmeans* in the Statistics and Machine Learning Toolbox™ which by default minimizes the sum of squared Euclidean distances to find the best number of clusters, *k*. LMS detection thresholds were selected as input features to start with since they clearly cluster the data points into separate groups (Figure S3). Results of using three features (LMS detection thresholds) or two features (LM thresholds) were compared. The number of clusters evaluated was set from 1 to 37 as there were 37 data points in total.

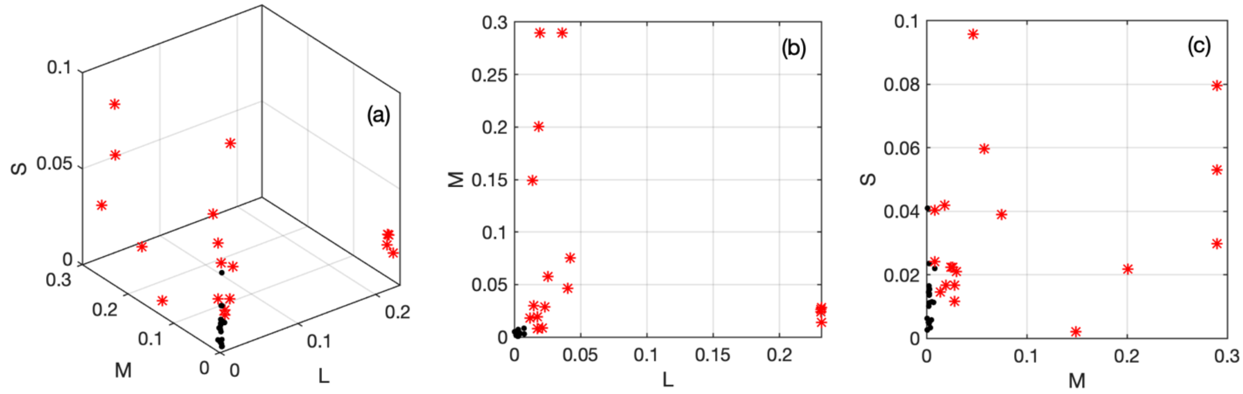


Figure S3. L-, M-, and S-cone detection thresholds in (a) LMS 3d view (b) M vs. L 2d view and (c) S vs. M 2d view. CN and CVD participants are denoted by black dots and red asterisks, respectively.

As in the elbow plot (Figure S4a), the sum of the within-cluster squared Euclidean distance versus number-of-cluster curve reveals a clear elbow at k=3. In Figure S4b, k=3 has the highest silhouette value among the reasonable number of clusters while all the values are larger than 0 indicating that when k=3, data points in each cluster are tightly grouped and no data points were misclassified. Silhouette coefficients of all data points at k=3 are larger than 0.78, except for CVD# 18 who has a silhouette value of 0.0793. Therefore, with three features defined, the first-round classification achieved a best k of 3, with a mean silhouette value of 0.9294. Pseudo F statistics (between-cluster sum of squared distance/ within-cluster sum of squared distance) of each feature in each cluster are reported in Table S3 as an indicator of the contribution of each feature to each cluster. When defining only two features (LM thresholds), the elbow plot indicates an optimal *k* at 3; whereas based on the Silhouette plot, k=3 and 4 respectively have a mean Silhouette coefficient of 0.9475 and 0.9478. With two features, k-means algorithm was able to classify individuals to the same three clusters with high Silhouette coefficients as compared to the previous analysis (Figure S5).

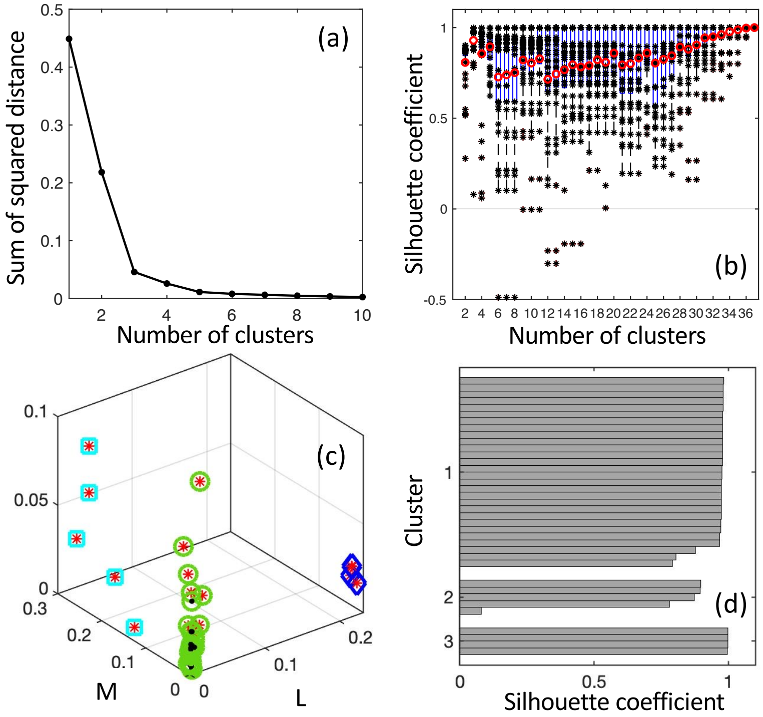


Figure S4. Step-one clustering with three input features (LMS). (a) Elbow plot with vertical and horizontal axes being sum of squared distance and number of clusters, showing a clear elbow of k=3. (b) Silhouette plot which plots silhouette coefficient versus number of clusters. (c) Data points clustered as three groups in the LMS detection threshold space. Representation of the data points is adopted from Figure S3a. The three groups are denoted in green circles, cyan squares, and blue diamonds surrounding the data points. (d) Silhouette coefficients for each data points when k=3.

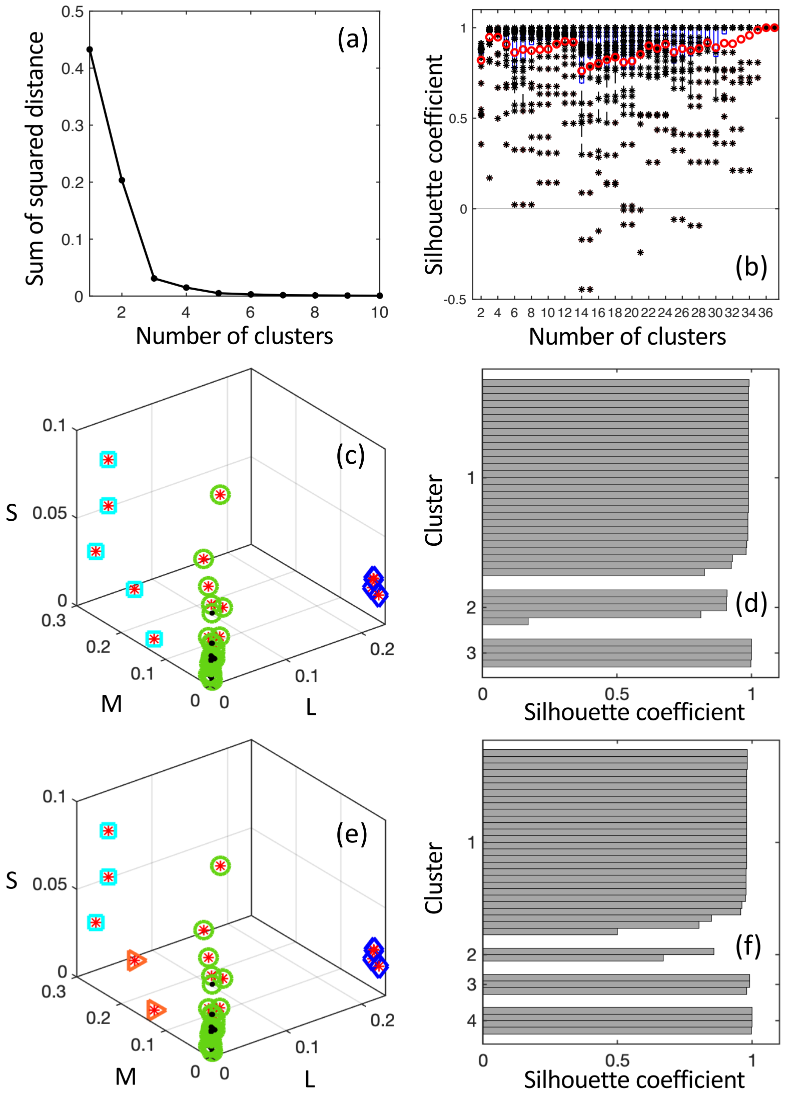


Figure S5. Step-one clustering with two input features (LM). (a) Elbow plot with vertical and horizontal axes being sum of squared distance and number of clusters, showing a clear elbow of k=3. (b) Silhouette plot which plots silhouette coefficient versus number of clusters. (c) Data points clustered as three groups in the LMS detection threshold space. Representation of the data points is adopted from Figure S3a. The three groups are denoted in green circles, cyan squares, and blue diamonds. (d) Silhouette coefficients for each data points when k=3. (e) is similar to (c) but data points are clustered into four groups. The fourth group is denoted by orange triangle. (f) Silhouette coefficients for each data points when k=4.

| **3 features (LMS) 3 clusters** | | | |
|---|---|---|---|
| | Cluster1(green circles; n=28) | Cluster2(cyan squares; n=5) | Cluster3(blue diamonds; n=4) |
| Feature1:L | 23.3121 (71.65%) | 17.8675 (86.83%) | 6.8992*10^7 (100%) |
| Feature2:M | 9.1971 (28.27%) | 2.6351 (12.81%) | 61.6367 (0%) |
| Feature3:S | 0.0277 (0.09%) | 0.0747 (0%) | 1.4007 (0%) |
| **2 features (LM) 3 clusters** | | | |
| | Cluster1(green circles; n=4) | Cluster2(cyan squares; n=28) | Cluster3(blue diamonds; n=5) |
| Feature1:L | 6.8992*10^7 (100%) | 23.3121 (71.71%) | 17.8675 (87.15%) |

| | | | |
|---|---|---|---|
| Feature2:M | 61.6367 (0%) | 9.1971 (28.29%) | 2.6351 (12.85%) |

| **2 features (LM) 4 clusters** | | | | |
|---|---|---|---|---|
| | Cluster1(green circle; n=28) | Cluster2(orange triangle; n=2) | Cluster3(cyan square; n=3) | Cluster4(blue diamonds; n=4) |
| Feature1:L | 9.6587 (44.72%) | 91.5145 (99.33%) | 5.9588 (0%) | 5.7382*10^7 (100%) |
| Feature2:M | 11.9414 (55.28%) | 0.6143 (0.67%) | ∞ (100%) | 87.1094 (0%) |

Table S3. Pseudo F statistics defined as between-cluster sum of squared distance divided by within-cluster sum of squared distance. Values in the parentheses indicate percentage contribution of the features to each cluster.

Common characteristics shared by each of the clusters were scrutinized for all classification possibilities. In the cases of k=3, five observers were classified in cluster 2 (CVD#2,3,4,6,18) shown as cyan open squares, and all were self-reported CVD and classified as medium/strong Deutan by HRR and FM100. Four observers classified in cluster 3 (CVD#1,5,10,14), shown as open blue diamonds, were all self-reported CVD and classified as medium or strong Protan by HRR and 100hue. Cluster 1, represented by open green circles, contains all normal color participants and 9 self-reported CVD. These 9 CVD participants have much lower L,M thresholds compared to CVDs in the other two clusters, and were categorized as either protan or deutan with different severity by HRR and 100hue. These 9 participants are likely protanomaly or deuteranomaly with the commonly used color vision categories for CVD type. When k=4, the only difference is that the members in cluster 2 at k=3 were divided into two clusters: CVD#2,4,6 who have the highest M-cone thresholds (likely strong deutan) and CVD#3,18 whose M-cone thresholds were in between two other clusters (likely medium deutan).

A second-round clustering was completed to further differentiate potential anomalous trichromats (AT) and CN participants (only the individuals grouped in the cluster denoted by green circles in Figure S4, S5). Based on the first-round classification results, the 5 Deutan participants reveal "M"-shape pattern with Y and B being the highest in the discrimination threshold plots (Figure 2), and the 5 Protan participants reveal "N"-shape pattern with Y and M being the highest. Therefore, thresholds of these three hues (Y, B, M) are of interest in the second-round classification. Given that low- and high-saturation levels reveal similar patterns and low-saturation thresholds show more distinct patterns, here we only focus on low-saturation hue discrimination thresholds. Together with LMS detection thresholds, six input features were used. In Figure S6a, a clear elbow cannot be obtained solely based on the elbow plot, and in Figure S6b, the silhouette plot suggests a best k value of 4. Cluster 1 perfectly recognized all 19 CNs. Three CVDs were assigned to cluster 3 (CVD#:8,11,13), by HRR and FM100, two of which were classified as mild Deutan (CVD#8) and strong Deutan (CVD#13), and one with mild red-green deficiency (CVD#11) received contradict diagnoses. Only one of the 3 CVDs assigned to cluster 2 (CVD#:12,15,17) was classified as strong Protan (CVD#12), and the other two did not receive consistent diagnosis by HRR and FM100 in the test session. However, CVD#12 was classified by HRR as medium deutan in the retest session, which we consider an inconsistent diagnosis. In cluster 4, two of the three CVDs were classified as medium (CVD#16) and strong protan (CVD#9), and one (CVD#7) did not receive consistent diagnosis.

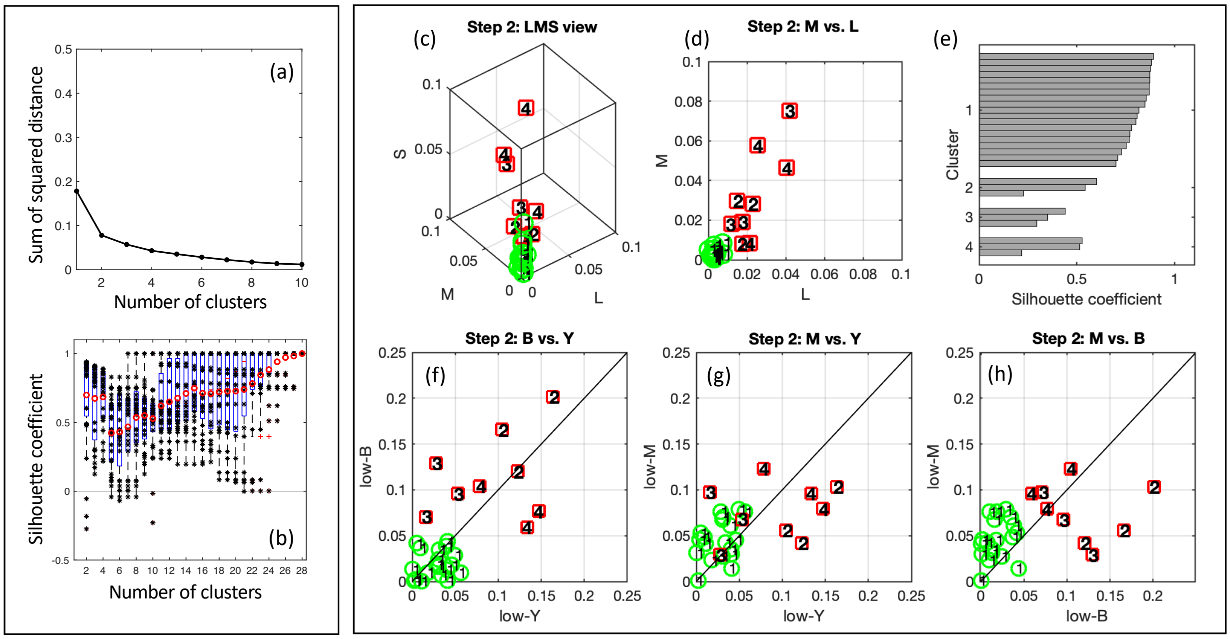


Figure S6. Step-two clustering with six input features (LMS,YBM). (a) Elbow plot. Similar to Figure S4a but without a clear elbow. (b) Silhouette plot. Similar to Figure S4b. (c) Data points clustered into four groups in the LMS detection threshold space. CN and CVD individuals are denoted by green circles and red squares, respectively. The four groups are denoted in numbers (1 through 4). (d) Top view of (c). (e) Silhouette coefficients for each data points at k=4. (f)-(h) the same set of data points plotted in three other views: B vs. Y, M vs. Y, and M vs. B. Panels (c),(d),(f),(g),(h) are also included in Figure 3.

| | Cluster1(Normal;n=19) | Cluster2(UnknownAT;n=3) | Cluster3(Deutan;n=3) | Cluster4(Protan;n=3) |
|---|---|---|---|---|
| Feature1:L | <u>25.3583 (33.47%)</u> | 0.0009 (0.03%) | 0.0358 (1.76%) | <u>0.3658 (17.94%)</u> |
| Feature2:M | <u>37.9397 (50.07%)</u> | 0.0196 (0.56%) | 0.0484 (2.39%) | 0.0767 (3.76%) |
| Feature3:S | 1.5404 (2.03%) | <u>1.3810 (39.64%)</u> | 0.0001 (0.01%) | <u>0.4542 (22.27%)</u> |
| Feature4:Y | 2.6946 (3.56%) | <u>1.0258 (29.45%)</u> | <u>1.9112 (94.21%)</u> | <u>0.4546 (22.29%)</u> |
| Feature5:B | <u>7.8532 (10.37%)</u> | <u>1.0548 (30.28%)</u> | 0.0273 (1.34%) | 0.0648 (3.18%) |
| Feature6:M | 0.3799 (0.50%) | 0.0014 (0.04%) | 0.0060 (0.29%) | <u>0.6232 (30.56%)</u> |

Table S4. Pseudo F statistics at k=4. Values in the parentheses indicate percentage contribution of the features in each cluster. Contributions larger than 10% from features are underlined.

***Bland-Altman analysis***

Test-retest differences are presumed to be normally distributed. In Figure S7,S8, mean test-retest differences or bias ($\bar{d}$) of all scores and thresholds (thick black horizontal lines) are close to zero (dotted black horizontal lines) and within the 95% confidence interval (CI) range of $\bar{d}$ except for FM100 total error score (TES) which is slightly outside of the range of 95% CI. The upper and lower 95% limits of agreement (LoA) were calculated as $\bar{d}$±1.96×standard deviation of the difference ($s$). CVD data points spread out more than CN signifying larger test-retest difference in scores or thresholds for the CVD group in general. At lease 20/21 data points in each panel fall within the outer 95% confidence limits of LoAs. Best linear fits between $\bar{d}$ and means are shown in red dashed lines, all of which have $p$ values larger than 0.01. Systematic learning effect and bias were not found for all tasks. The coefficients of repeatability (COR) were also reported in Table S5.

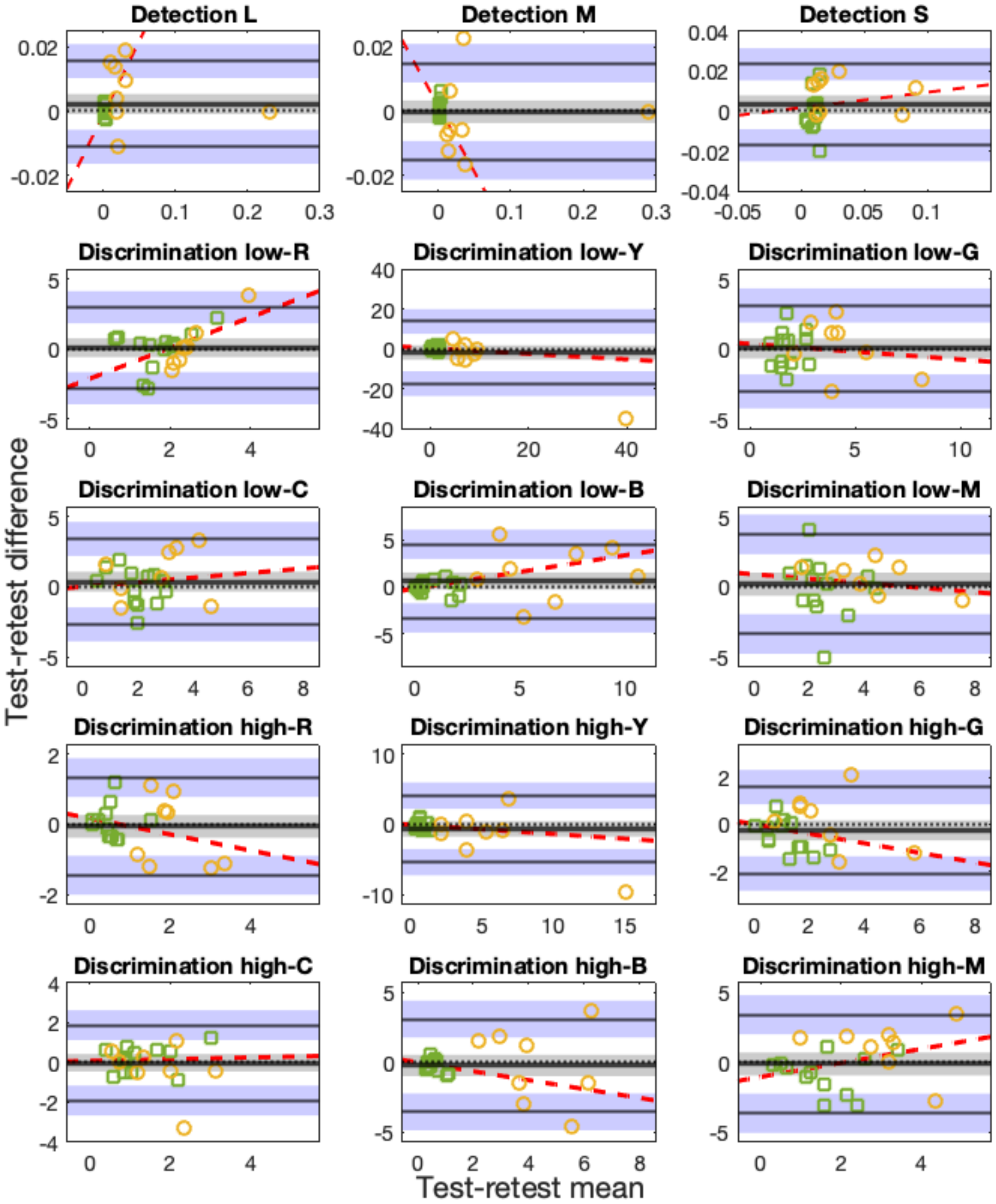


Figure S7: Bland-Altman plots for FInD Color detection L-, M-, and S-cone isolating thresholds and FInD Color discrimination low and high saturated red (R), yellow (Y), green (G), cyan (C), blue (B), magenta (M) thresholds in degree for 13 CN (green squares) and 8 CVD (orange circles) participants. The thicker solid black line in the middle indicates mean of the test-retest differences ($\bar{d}$) and the middle dotted black line indicates a difference of 0 as a reference. These two lines overlap in some panels. Limits of agreement (LoA) lines are depicted as upper and lower black lines. Grey and blue zones are 95%-CI ranges of $\bar{d}$ and LoAs, respectively, indicating precision of the estimates. Best linear fit is plotted in dashed red line. Note that scales are selected for each panel to best show data points.

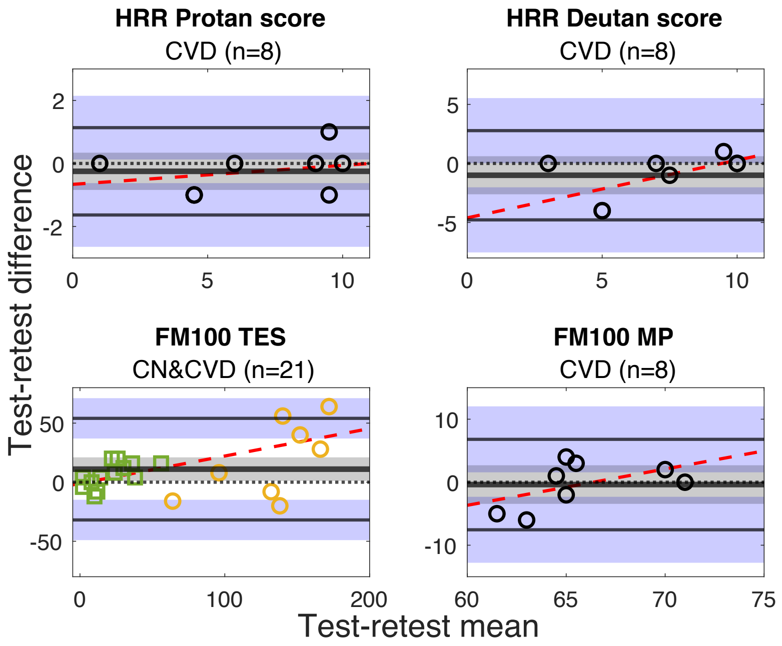


Figure S8: Bland-Altman plots for HRR Protan and Deutan scores, and FM100 total error scores (TES) and right-side middle-points (MP). Circles represent CVD data and green squares represent CN data.

| Statistics | *n* | $\bar{d}$ *(p)* | *s* | *95%CI of* $\bar{d}$ | *95%CI of LoAs* | *Slope (p)* | *COR* |
|---|---|---|---|---|---|---|---|
| HRR Protan | 8 | -0.25 (0.351) | 0.707 | ±0.591 | ±1.011 | 0.060 (0.380) | 1.482 |
| HRR Deutan | 8 | -1 (0.186) | 1.927 | ±1.611 | ±2.755 | 0.488 (0.074) | 4.320 |
| FM100 TES | 21 | 11.048 (0.032) | 21.942 | ±9.988 | ±17.079 | 0.232 (0.860) | 48.393 |
| FM100 MP | 8 | -0.375 (0.780) | 3.662 | ±3.062 | ±5.235 | 0.576 (0.222) | 7.221 |
| FInD detection L | 21 | 0.002 (0.148) | 0.007 | ±0.003 | ±0.005 | 0.459 (0.314) | 0.014 |
| FInD detection M | 21 | -0.0002 (0.921) | 0.008 | ±0.004 | ±0.006 | -0.411 (0.179) | 0.015 |
| FInD detection S | 21 | 0.003 (0.172) | 0.010 | ±0.005 | ±0.008 | 0.077 (0.592) | 0.021 |
| FInD discrimination low-R | 21 | 0.089 (0.785) | 1.479 | ±0.673 | ±1.151 | 1.099 (0.023) | 2.904 |
| FInD discrimination low-Y | 21 | -1.761 (0.322) | 7.940 | ±3.614 | ±6.180 | -0.140 (0.503) | 15.958 |
| FInD discrimination low-G | 21 | 0.043 (0.902) | 1.570 | ±0.715 | ±1.222 | -0.112 (0.623) | 3.078 |
| FInD discrimination low-C | 21 | 0.374 (0.283) | 1.554 | ±0.707 | ±1.210 | 0.993 (0.707) | 3.137 |
| FInD discrimination low-B | 21 | 0.623 (0.171) | 2.007 | ±0.914 | ±1.562 | 0.357 (0.651) | 4.128 |
| FInD discrimination low-M | 21 | 0.195 (0.627) | 1.811 | ±0.825 | ±1.410 | -0.160 (0.294) | 3.572 |
| FInD discrimination high-R | 21 | -0.058 (0.713) | 0.720 | ±0.323 | ±0.552 | -0.229 (0.520) | 1.395 |
| FInD discrimination high-Y | 21 | -0.642 (0.238) | 2.419 | ±1.101 | ±1.883 | -0.133 (0.892) | 4.913 |
| FInD discrimination high-G | 21 | -0.255 (0.230) | 0.943 | ±0.429 | ±0.734 | -0.204 (0.981) | 1.918 |

| | | | | | | | |
|---|---|---|---|---|---|---|---|
| FInD discrimination high-C | 21 | -0.026 (0.901) | 0.962 | ±0.438 | ±0.749 | 0.043 (0.847) | 1.887 |
| FInD discrimination high-B | 21 | -0.230 (0.541) | 1.695 | ±0.772 | ±1.319 | -0.317 (0.964) | 3.354 |
| FInD discrimination high-M | 21 | -0.125 (0.754) | 1.800 | ±0.819 | ±1.401 | 0.502 (0.230) | 3.537 |

Table S5: Statistics of the Bland-Altman analysis results: sample size (*n*), mean test-retest difference or bias ($\bar{d}$) and its *p* value, standard deviation of the differences (*s*), 95% confidence interval (CI) of $\bar{d}$, 95%CI of limits of agreement (*LoAs*), slope of the best fit line and its *p* value, and coefficient of repeatability (*COR*).

#### *Cone isolating directions for the display used*

Here we show cone isolating directions calculated based on Stockman-Sharpe cone fundamentals(1, 2) for the display employed (LG 32UD59-B). We measured the spectra of the display with a Photoresearch PR670 Spectroradiometer (Chatsworth, CA, USA), and plotted the curves in Figure S9. Calculated cone isolating directions are summarized in Table S6.

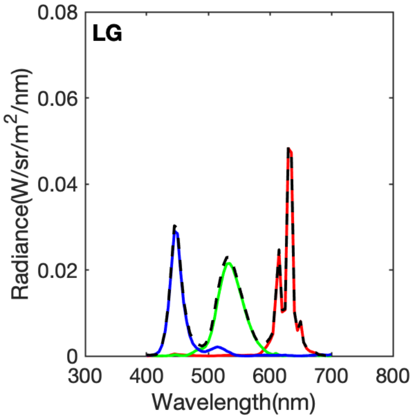


Figure S9. Spectra profiles of displays measured. Red (right distributions), green (middle distributions), and blue (left distributions) primary spectra are plotted as red, green, and blue curves respectively. White spectra, the summation of the three primaries, are represented by black dashed curves.

| Type | LED | | |
|---|---|---|---|
| | R | G | B |
| L | 1 | 0.44404 | 0.49532 |
| M | 0 | 0.75974 | 0.49413 |
| S | 0.53668 | 0.43073 | 1 |

Table S6: Cone isolating directions in RGB unit for the experimental display with values ranging from 0 to 1.

#### *Luminance artifact analysis*

Here we analyze the magnitude of the luminance artifacts induced in the FInD Color discrimination task. Figure S10 top middle panel shows that the low saturation color blobs selected from the HSV plane can generate a luminance difference as large as 39 cd/m$^2$. With a threshold of 5° hue angle (thresholds of CN group are below 5°), luminance differences between the color blobs are within the range of ±3 cd/m$^2$ (Figure S10 top right panel). We also measured discrimination thresholds of 4 color normal participants to achromatic blobs using FInD algorithm at luminance levels that are close to the six HSV hue angles we used. Results shown that the lowest possible mean luminance threshold is about 7.2° in low-saturation discrimination task (Table S7, hue=M) which is larger than the 5° chromatic discrimination threshold, suggesting that it was the chromatic signals that were used by the participants to reach the discrimination thresholds and that luminance artefacts do not affect the present results. However, the high-saturation discrimination stimuli might induce detectable luminance artifacts for red, cyan, and magenta because achromatic discrimination thresholds along three axes are close to the 5° chromatic discrimination threshold limit (Table S7: R, C, M). This might explain the pattern difference observed between low- and high-saturation discrimination thresholds, as low-saturation stimuli better revealed the real chromatic discrimination thresholds and produced more distinct patterns, whereas high-saturation stimuli might reduce the differences between CN and CVD groups. Note that our diagnostic analyses were based on only low-saturation discrimination thresholds.

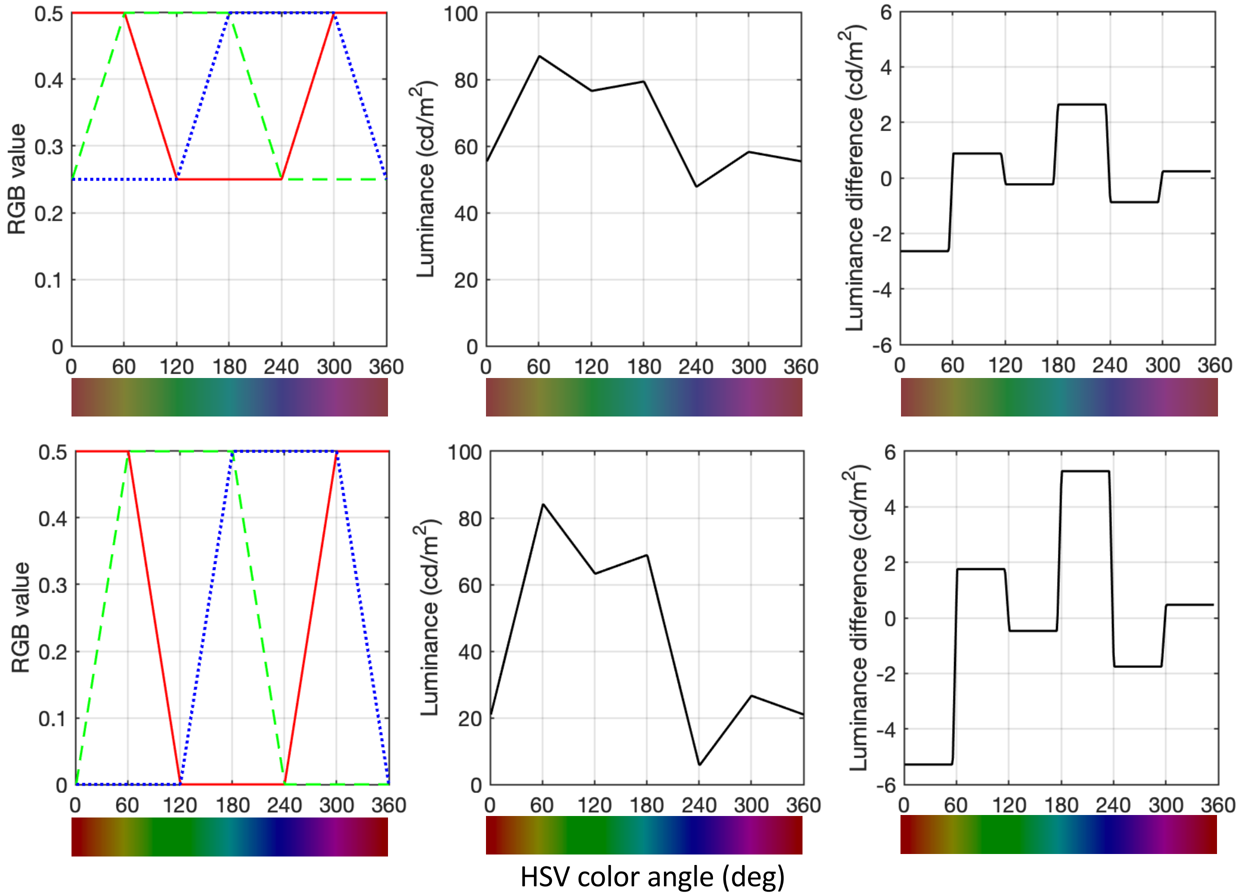


Figure S10. Luminance artifact analysis for FInD Color discrimination task. Upper panels and lower panels are for low- and high-saturation stimuli, respectively. Left panels: RGB values of HSV color angles. Red, green, and blue channel values are shown in red solid, green dashed, and blue dotted lines. Middle panels: luminance of HSV color angles. Right panels: luminance differences of HSV color angles with a threshold angle of 5°.

| Hue (angle) | R (0°) | Y (60°) | G (120°) | B (180°) | C (240°) | M (300°) |
|---|---|---|---|---|---|---|
| Low saturation | 25.08 (±3.87) | 16.48 (±4.99) | 7.67 (±0.11) | 19.03 (±8.57) | 19.30 (±2.12) | 7.50 (±0.30) |

| | | | | | | |
|---|---|---|---|---|---|---|
| High saturation | 5.45 (±0.89) | 16.39 (±2.75) | 14.65 (±1.27) | 19.00 (±8.30) | 5.21 (±1.53) | 5.48 (±1.56) |

Table S7. Mean (±standard deviation) achromatic discrimination threshold in cd/m$^2$ of four CN participants.

***FInD psychometric function parameters***

The detection and discrimination thresholds were estimated by fitting a psychometric function (Eq.1) to the responses. The goodness of fit of the fitted curve, which depends on the parameter values, determines the quality of estimated thresholds. This psychometric function is constrained by four factors: weight for each response, starting values, lower bounds, and upper bounds for the three parameters (false alarm rate, threshold, slope). The response weights were inverse of binomial standard deviation for each test level. Parameters values used are summarized in Table S8. The psychometric function was properly fitted to most of the data (e.g. CVD1 M,S and CVD2 L,S in Figure S11), but not all. The upper left panel in Figure S11 is an example of a poorly fitted curve, where the function does not fit to seemingly random responses. This situation sometimes occurs when the defective cone type is tested, thus we assign the maximal level tested to it as the threshold. CVD 2 middle panel in Figure S11 shows another case when the defective M cone was tested. The CVD participant was not able to see any of the M-cone isolating blobs and did not respond “yes” for all cells. In this case, a maximal test level is assigned as threshold as well.

| **Parameters** | **starting value** | **lower bound** | **upper bound** |
|---|---|---|---|
| False Alarm Rate | 0.01 | 0.01 | 0.75 |
| Threshold | Mean of test levels | Minimal test level | Maximal test level |
| Slope | 2 | Minimal difference between test levels | 4 |

Table S8: Parameters values.

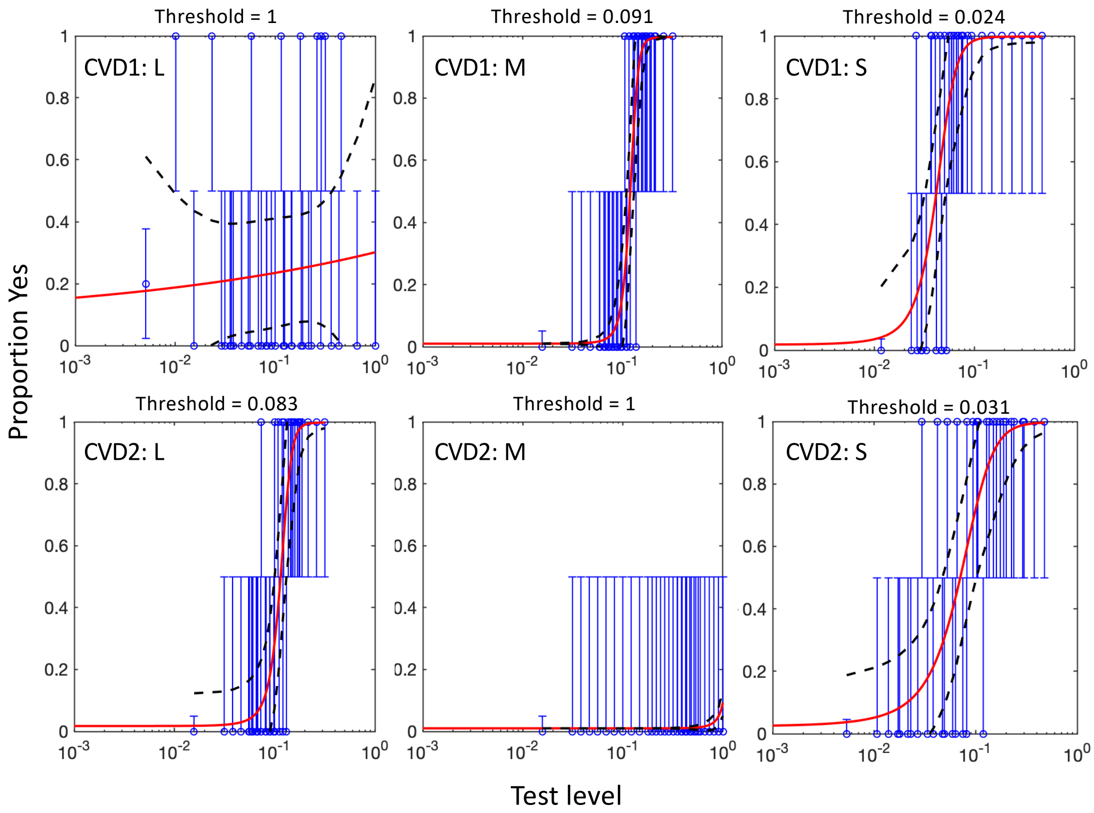


Figure S11. Example psychometric functions (red curves). Yes proportion data are shown in blue lines, and 95% confidence intervals are shown in black dashed curves. The separate data point in each panel on the left of the responses indicates the corresponding false alarm rate. Each row shows detection data of a single participant, and the columns from left to right show L, M, and S detection data. Thresholds reported above the panels range from 0 to 1, which have not been weighted by cone contrast vector length, to better exhibit the relationship between fitted curves and the estimated thresholds. CVD1 is a protan and CVD2 is a deutan.

***FInD Color discrimination task with an equiluminant color plane***

Six CVDs and 14 CNs have completed FInD Color discrimination task with colors chosen from an equiluminant plane instead of the HSV color space. The equiluminant plane contains 4 primary color directions, red-green axis (L-M, M-L) and purple-yellow axis (S+,S-), with red (R), purple (P), green (G), and yellow (Y) being 0°, 90°, 180°, 270°, respectively. The four intermediate angles were also tested (RP, PG, GY, YR). The calculation of the equiluminant plane is the same as in Eskew, Newton (3)(see *2.4. the equiluminant plane*). Discrimination thresholds of all participants are shown in Figure 2(c). T-tests were done for comparing whether, at each color axis, the threshold of one CVD participant is different from those of CNs. P-values are shown in Table S9.

| CVD# | Type | R | RP | P | PG | G | GY | Y | YR |
|---|---|---|---|---|---|---|---|---|---|
| 2 | Deuteranope | *** | 0.875 | *** | *** | *** | * | *** | *** |
| 5 | Protanope | *** | ** | *** | 0.427 | *** | ** | *** | 0.063 |
| 11 | Deuteranomaly | *** | 0.527 | *** | *** | *** | 0.052 | *** | 0.810 |
| 14 | Protanope | * | 0.084 | *** | * | *** | ** | *** | 0.129 |

| | | | | | | | | | |
|---|---|---|---|---|---|---|---|---|---|
| 15 | Deuteranomaly | *** | ** | *** | *** | *** | 0.084 | *** | ** |
| 17 | Protanomaly | *** | *** | *** | ** | *** | ** | *** | 0.529 |

Table S9. P-values for discrimination task with the equiluminant plane. $p < 0.001$, $p < 0.01$, and $p < 0.05$ are represented by ***, **, and *, respectively.

| CVD# | R | Y | G | C | B | M |
|---|---|---|---|---|---|---|
| 1 | 0.292 | *** | *** | *** | *** | *** |
| 2 | *** | *** | *** | *** | *** | *** |
| 3 | *** | *** | *** | *** | *** | *** |
| 4 | 0.057 | *** | *** | 0.090 | *** | *** |
| 5 | * | *** | *** | *** | *** | *** |
| 6 | *** | *** | *** | *** | *** | 0.055 |
| 7 | 0.104 | *** | 0.070 | ** | *** | *** |
| 8 | 0.331 | *** | * | 0.185 | *** | *** |
| 9 | *** | *** | *** | *** | *** | *** |
| 10 | * | *** | *** | 0.117 | *** | *** |
| 11 | 0.668 | *** | *** | 0.353 | *** | *** |
| 12 | 0.417 | *** | *** | *** | *** | 0.159 |
| 13 | *** | *** | *** | ** | *** | *** |
| 14 | 0.083 | *** | *** | *** | *** | ** |
| 15 | *** | *** | *** | *** | *** | *** |
| 16 | ** | *** | *** | *** | *** | *** |
| 17 | 0.264 | *** | *** | *** | *** | 0.624 |
| 18 | 0.463 | *** | *** | *** | *** | *** |

Table S10. P-values for discrimination task with the HSV plane. $p < 0.001$, $p < 0.01$, and $p < 0.05$ are represented by ***, **, and *, respectively.

***Rayleigh color matching task***

Participants wore their own correction lenses for the anomaloscope testing and used their preferred eye. Matching range was determined by asking the participants to indicate the leftmost (highest anomalous quotient) and rightmost matches (lowest anomalous quotient). The experimenter selected a set of stimuli across a large range of color pairs, and the participant was asked to report color appearance of the stimulus and whether the top and bottom colors are matched in brightness and hue until the colors appear to be matched. The resulting anomalous quotient (AQ) ranges and for the CVD participants are reported in Table S11. Severity categories are simple, extreme, or dichromat.

| CVD# | Classification | Severity | AQ range | | Matching range | |
|---|---|---|---|---|---|---|
| | | | Min. | Max. | Min. | Max. |
| 2 | Deuteranopia | dichromat | 0.09 | 16.1 | 5.1 (22.7) | 68.2 (13.3) |

| | | | | | | |
|---|---|---|---|---|---|---|
| 5 | Protanopia | dichromat | 0.02 | Infinity | 0 (41.2) | 71.8 (2.4) |
| 11 | Deuteranomaly | simple | 2.2 | 3.6 | 18.4 (15.7) | 25.9 (10.2) |
| 14 | Protanopia | dichromat | 0.13 | Infinity | 0 (32.6) | 66.1 (4.4) |
| 15 | Deuteranomaly | simple | 2.6 | 3.82 | 17.6 (11.8) | 23.2 (13.6) |
| 17 | Protanomaly | simple | 0.22 | 0.32 | 57.6 (6.3) | 62 (5.5) |

Table S11. Anomalous quotient (AQ) ranges and matching ranges for Rayleigh matches. According to the anomaloscope user's manual (Oculus, Germany), AQ ranging from 0.7 to 1.4 indicate normal matching zone; AQ ranging from less than 0.7 to 0.1 indicate Protanomaly; AQ ranging from larger than 1.4 (mostly larger than 2.0) to infinity indicate Deuteranomaly; AQ up to infinity or down to 0 or including the normal mid-match indicate extreme anomaly. The full range of the mixtures is acceptable for a dichromat. Numbers in the parentheses represent the corresponding reference light (yellow light setting).

***References***

1. Stockman A, Sharpe LT, Fach C. The spectral sensitivity of the human short-wavelength sensitive cones derived from thresholds and color matches. Vision research. 1999;39(17):2901-27.
2. Stockman A, Sharpe LT. The spectral sensitivities of the middle- and long-wavelength-sensitive cones derived from measurements in observers of known genotype. Vision Research. 2000;40(13):1711-37.
3. Eskew RT, Jr., Newton JR, Giulianini F. Chromatic detection and discrimination analyzed by a Bayesian classifier. Vision Research. 2001;41(7):893-909.
